# Disentangling Confounders from Pathology in Long-COVID Trajectory Prediction for Women: An Interpretable Large-Language-Model Approach

**DOI:** 10.64898/2026.06.10.26355420

**Authors:** Jing Wang, Zorina Galis, Tong Zhang, Yiming Luo, Amar Sra, Xing Niu, Jie Shen, Qiaomin Xie, Jeremy C. Weiss

## Abstract

**Objective:** Post-acute sequelae of SARS-CoV-2 infection (PASC, “Long COVID”) disproportionately affects women, in whom hallmark symptoms—insomnia, fatigue, palpitations, cognitive difficulty—overlap with comorbidities and hormonal transitions such as menopause. This diagnostic overlap is a confounding problem: models that forecast future symptom severity risk attributing baseline physiological noise to viral pathology. We ask whether an interpretable, causally disentangled language model can separate true pathological signal from such confounders while remaining competitive with strong predictors of future PASC severity.

**Materials and methods:** Retrospective cohort of 1,155 adult women (median age 61) from the NIH RECOVER program, combining static clinical profiles, longitudinal symptom surveys, and four weeks of mean-aggregated consumer-wearable physiology (heart rate, sleep, activity). We render each patient as a natural-language clinical narrative and fine-tune a small open-weight language model (Qwen2.5-0.5B, LoRA) with an attention-based *disentanglement* layer that gates the latent state into a causal and a confounder component, trained with an environment-mixing InfoNCE objective. We predict the PASC index at 3-, 6-, and 9-month horizons and benchmark against last-value carry-forward, Lasso, Ridge, gradient-boosted trees (XGBoost), a deep MLP, a tabular ResNet, and a self-attention network, over 20 stratified resamples with paired significance tests. We further stratify by trajectory phenotype (Protected / Responder / Refractory).

**Results:** Long-COVID severity is strongly autocorrelated, so last-value carry-forward is a hard reference and is the most accurate method on the full cohort (MAE 3.02*/*1.99*/*1.52 at 3/6/9 months). Among *learned* models the LLM regressor had the lowest MAE at every horizon (e.g. 3.11 vs. XGBoost 3.57 at 3 months; paired *p ≤* 0.01). In the Responder phenotype— patients whose trajectories actually move—the LLM was the most accurate method overall at 3 and 6 months (MAE 4.72, 4.06), though its advantage over carry-forward was not statistically significant (*p* = 0.70, 0.29). The disentanglement layer assigned maximal saliency to direct pathology tokens (*breathlessness, malaise*; 1.00) while suppressing confounders (*menopause, diabetes*; < 0.27) and linguistic filler (< 0.17).

**Conclusion:** For static, slowly evolving patients a simple carry-forward forecast is hard to beat and should be the reference any PASC model is judged against. The value of a learned, disentangled model is (i) better accuracy where trajectories are dynamic and (ii) an interpretable, “clinically honest” attribution that down-weights confounders such as menopause—reducing the risk of misattributing baseline physiology to Long COVID.

**Author summary:** Long COVID is more common and often more severe in women, but many of its symptoms look like other common conditions or like the normal changes of menopause. When a computer model tries to predict how a patient’s symptoms will evolve, it can be fooled into blaming Long COVID for what is really background physiology. We built a model based on a small language model that reads a written summary of each patient—their history, comorbidities, and a month of wearable-device data—and is explicitly trained to separate “true disease signal” from “background noise.” We tested it against standard predictors at 3, 6, and 9 months. We report an finding that is easy to overlook: because Long COVID severity changes slowly, simply assuming a patient’s next score equals their last score is very accurate and hard to beat for stable patients. Our model’s advantage appears where it matters clinically—patients whose symptoms are actually changing—and, importantly, the model shows *which words drove its prediction*, correctly emphasizing symptoms like breathlessness while down-weighting confounders like menopause. We argue this interpretability, not a small accuracy gain, is the real contribution.

## 1 Introduction

Persistent, multisystem symptoms following SARS-CoV-2 infection—post-acute sequelae of SARS-CoV-2 (PASC), or Long COVID—have become a global public-health burden, with profound fatigue, cognitive impairment (“brain fog”), dysautonomia, and respiratory complaints lasting months to years after viral clearance [1–3]. The burden is not distributed evenly: women, and in particular women in midlife, carry a disproportionate share of risk, with reports of an approximately 40% higher PASC risk than age-matched men and an apparent interaction with the menopausal transition [4, 5].

This sex disparity creates a specific methodological hazard. Many cardinal PASC symptoms— insomnia, palpitations, joint pain, vasomotor instability, fatigue—are also cardinal symptoms of menopause and of common cardiometabolic and mental-health comorbidities [6]. A predictive model trained on such data can achieve a good error metric while learning the *wrong* thing: attributing a patient’s elevated future symptom burden to viral pathology when it is in fact driven by a confounder. Classical causal-inference work has long emphasized that failing to adjust for such confounding biases downstream conclusions [7, 8]. In a clinical deployment, a model that conflates menopause with active Long COVID would systematically over-flag a large subpopulation, eroding trust and clinical utility.

Consumer wearables offer an objective, longitudinal complement to self-report: heart-rate variability, sleep architecture, and activity proxies can capture autonomic and behavioral shifts associated with PASC [9, 10]. But wearable streams are high-dimensional and noisy, and the same signals (e.g. resting heart rate, sleep fragmentation) respond to both PASC and ordinary aging.

Large language models (LLMs) can fuse heterogeneous clinical narratives and have shown promise as flexible clinical encoders [11, 12]; yet a vanilla LLM optimizes for predictive correlation and will happily exploit confounders if they reduce loss.

### Contributions

We take an *interpretability-first*, honestly benchmarked position.

1. We build a causally disentangled LLM regressor that gates its latent representation into a causal and a confounder component and is trained with an environment-mixing contrastive objective, so that the readout depends on pathological signal rather than patient background.
2. We forecast PASC severity at 3-, 6-, and 9-month horizons and benchmark against a full suite of baselines—crucially including a last-value carry-forward reference and gradient-boosted trees—with paired statistical tests over 20 resamples. We report the result: for the full cohort carry-forward is the strongest method, and the learned models’ value is conditional.
3. We show that the model’s value is (a) accuracy in the dynamic *Responder* phenotype and (b) a stable, clinically sensible attribution that maximizes saliency on pathology tokens and suppresses confounders such as menopause and diabetes—a property no error metric captures.

## 2 Materials and methods

### 2.1 Study population and data source

We used data from the NIH RECOVER (Researching COVID to Enhance Recovery) program,^1^ a national prospective effort to characterize the long-term effects of COVID-19. Our analytic cohort comprised 1,155 adult women for whom we could link three modalities: (i) static demographic and clinical profiles, (ii) repeated PASC symptom surveys, and (iii) consumer-wearable physiology. Baseline characteristics are summarized in Table 1. The cohort is predominantly midlife-to-older (median age 61, IQR 56–67) and racially/ethnically diverse, with a high prevalence of comorbidities that overlap with PASC: 59.9% reported heart conditions, 27.5% sleep disorders, 31.4% mental-health conditions, and 68.9% had reached menopause—the central confounder of interest. Figure 1 shows the comorbidity distribution.

**Table 1:**
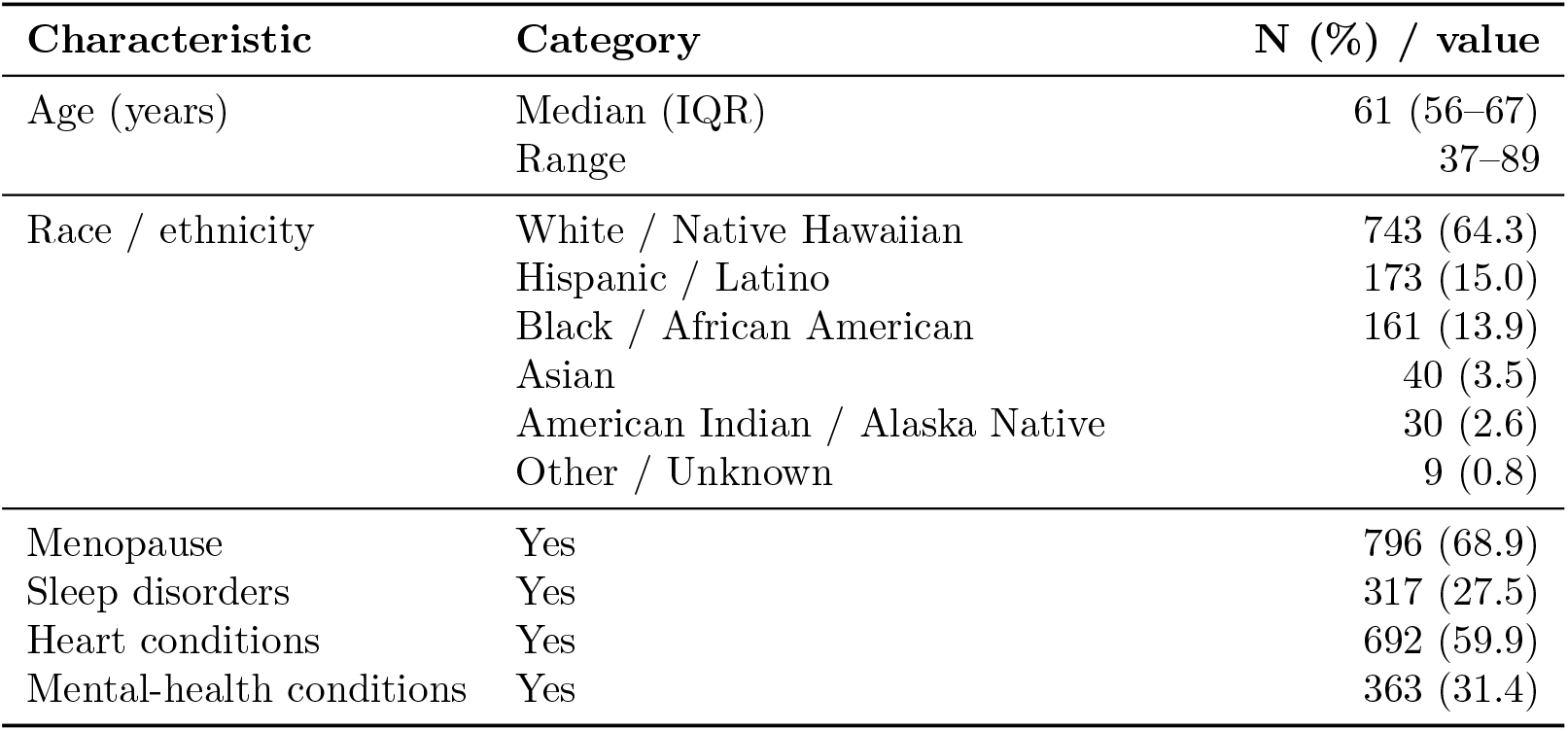
Baseline demographic and clinical characteristics (N = 1,155).

**Figure 1:**
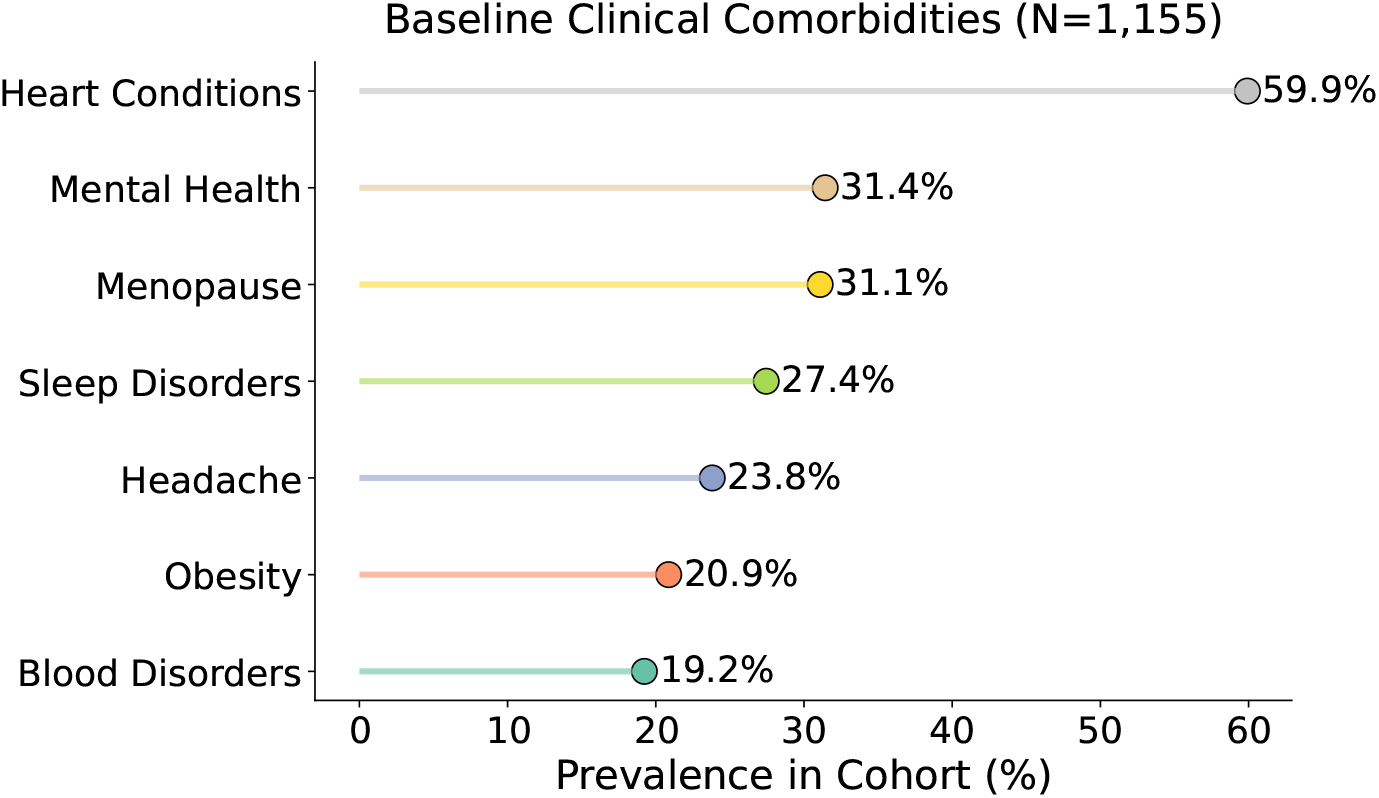
Prevalence of baseline comorbidities in the cohort. Several of the most prevalent conditions (cardiac, sleep, mental-health, menopause) share symptomatology with PASC, motivating the confounder-disentanglement objective.

### 2.2 Outcome, horizons, and trajectory phenotypes

The prediction target is the per-visit PASC symptom index (a 0–30 severity score derived from the Thaweethai *et al*. symptom set [2]). Each woman’s baseline visit is anchored to her published consecutive survey pair; we then take the survey nearest to +91, +183, and +274 days (within tolerance windows of 45–135, 105–300, and 180–420 days, respectively) to define the 3-, 6-, and 9-month targets. Because PASC follow-up is irregular, the retained sample shrinks with horizon— an attrition we report rather than hide: analytic *N* = 1090*/*615*/*546 at 3/6/9 months on the full cohort.

To probe *when* learning helps, we stratify the cohort into three trajectory phenotypes used throughout RECOVER analyses: **Protected** (consistently low/stable scores; low trajectory variance), **Responder** (dynamic, materially changing scores; high variance), and **Refractory** (persistently high scores). Empirically the phenotypes differ sharply in trajectory variability (Responder symptom-score SD *≈* 5.0 vs. Protected *≈* 1.4), which—as we show—is exactly the axis along which a learned model can add value over carry-forward. *(Exact phenotype thresholds are fixed by the RECOVER extraction script; see Data availability*.)

### 2.3 Wearable feature construction

For each horizon window we aggregated raw wearable streams (cardiac/respiratory rate, sleep architecture, activity intensity) into four equal time bins, yielding a fixed feature schema (*concept_Value*); missing channels were encoded with a sentinel (*−*1) matching the original gating. Wearable coverage is partial (with-wearable *N* = 173*/*88*/*86 at 3/6/9 months); the sentinel encoding lets the model use physiology when present without dropping patients when absent. *Caveat (stated for transparency):* for the longer horizons the wearable window overlaps the outcome survey, so those features are best read as *near-real-time monitoring* inputs rather than as strictly pre-baseline predictors; a strictly baseline-only variant is a planned sensitivity analysis (see Discussion).

### 2.4 Language-model regressor and the disentanglement variant

We use a single Qwen2.5-0.5B backbone [13] in two configurations, and we are explicit about which is used for which result. (i) The **LLM regressor** renders each patient as a structured natural-language narrative (static profile, historical PASC score, binned wearable values), encodes it with LoRA (rank 32, *α* = 64, *q, k, v, o* projections, dropout 0.1), and adds a learned residual gate on the historical PASC value before a clamped [0, 30] readout. This is the model evaluated across all 3/6/9-month horizons and cohorts in Table 2. (ii) The **causal-disentanglement variant** adds the attention-gating, environment-mixing, and InfoNCE machinery below; it is the model used for the interpretability/saliency analysis (Table 3, Fig. 3) and, in the present study, was evaluated at the 3-month horizon. Extending the disentanglement variant to all horizons is the most important follow-up (see Limitations). We describe the disentanglement mechanism next.

**Table 2:** Mean absolute error (PASC points; mean over resamples) by method, horizon, and cohort. **Bold** = best in column. “LLM regressor” is the narrative Qwen model (configuration i); “History” is last-value carry-forward. Among *learned* models the LLM regressor is best in every column. The causal-disentanglement variant (configuration ii) is used for the interpretability analysis and was evaluated at 3 months (3-month MAE 4.15 *±* 1.21 on the full cohort), where it trades raw error for the attribution properties in Table 3.

| Method | Full cohort |  |  | Responder phenotype |  |  |
| --- | --- | --- | --- | --- | --- | --- |
|  | 3m | 6m | 9m | 3m | 6m | 9m |
| History (carry-forward) | **3.02** | **1.99** | **1.52** | 4.82 | 4.36 | 4.50 |
| Lasso | 3.35 | 2.48 | 1.95 | 4.86 | 4.68 | 4.80 |
| Ridge | 3.53 | 2.74 | 2.18 | 5.13 | 5.26 | 5.39 |
| XGBoost | 3.57 | 2.67 | 2.18 | 5.21 | 5.12 | 5.56 |
| Deep MLP | 3.49 | 2.55 | 2.11 | 5.14 | 5.44 | 5.63 |
| ResNet | 3.60 | 2.57 | 2.11 | 5.23 | 5.12 | 6.06 |
| Attention | 5.49 | 4.22 | 3.48 | 5.50 | 5.18 | 5.54 |
| **LLM regressor (Ours)** | 3.11 | 1.98 | 1.66 | **4.72** | **4.06** | **4.45** |

**Table 3:** Causal-saliency attribution of representative tokens (normalized routing weight toward the causal head; 1.00 = exclusively causal, 0.00 = treated as noise).

| Category | Representative tokens | Max saliency | Interpretation |
| --- | --- | --- | --- |
| Active pathology | *breath, malaise, unrefreshing* | 1.00 | Direct pulmonary/systemic PASC signal. |
| Cognitive symptoms | *fog, brain, insomnia* | 0.54–0.74 | Hallmark neuro-PASC cluster. |
| Confounders | *menopause, diabetes* | < 0.27 | Down-weighted as non-causal background. |
| Administrative / filler | *that, to, by* | < 0.17 | Ignored as low-information tokens. |

Let **h** *∈* ℝ^*d*^ be the last-token hidden state. A *disentanglement attention layer* produces two scalar gates,

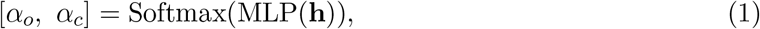

splitting **h** into a **causal** component **x**_*o*_ = *α*_*o*_**h** (invariant pathology, e.g. “malaise,” “breathlessness”) and a **confounder** component **x**_*c*_ = *α*_*c*_**h** (baseline noise, e.g. menopause status, diabetes, linguistic filler). To prevent reliance on patient-background spurious correlations, we apply *environment mixing*: a counterfactual representation combines patient *i*’s causal features with a randomly drawn patient *j*’s confounder features,

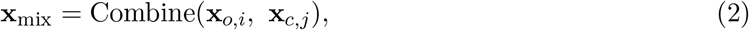

forcing the readout to predict from **x**_*o,i*_ regardless of the confounding environment. An InfoNCE contrastive term [14] aligns the mixed representation with the original to preserve semantic consistency,

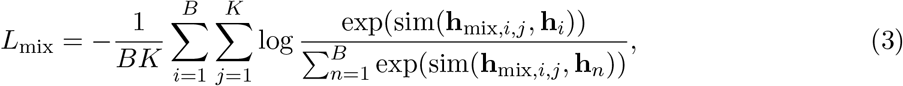

with cosine similarity sim(·), batch size *B*, and *K* negatives. Three heads—causal *f*_*o*_(**x**_*o*_), confounder *f*_*c*_(**x**_*c*_), and joint *f*_*co*_(**x**_mix_)—are supervised against the ground-truth score. The final prediction adds a learned residual gate on the historical PASC value and is clamped to [0, 30]. Training used AdamW (lr 2 *×* 10^*−*5^, weight decay 0.01), SmoothL1 loss, gradient clipping at 1.0, batch size 16, 10 epochs; inputs truncated/padded to 256 tokens (left padding). The open-weight model was fine-tuned and run entirely locally on AWS g5 instances (NVIDIA A10G GPUs) within the RECOVER-approved secure environment; local deployment satisfies the HIPAA and data-use-agreement constraints under which RECOVER data cannot be transmitted to external API endpoints.

### 2.5 Baselines, protocol, and statistics

We benchmarked against: **History** (last-value carry-forward of the patient’s most recent PASC score—the clinically obvious reference), **Lasso** and **Ridge** (CV-tuned) [15, 16], **XGBoost** [17], a **deep MLP** with a feature gate, a tabular **ResNet** [18], and a self-**attention** network [19]. Tabular models used 20 stratified 80/20 resamples; the LLM used 10 (compute-bound). All regression predictions were clamped to [0, 30]. We report MAE, RMSE, and MSE as mean*±*SD. For the headline comparisons we used paired tests across matched resamples (Wilcoxon signed-rank and paired *t*) between the LLM and the carry-forward reference, and between the LLM and XGBoost. Reporting follows the TRIPOD+AI checklist for prediction-model studies (Supplement).

## 3 Results

### 3.1 Carry-forward is the reference, and it is hard to beat on the full cohort

Table 2 reports MAE by method and horizon. Because PASC severity is strongly autocorrelated, last-value carry-forward (History) is the most accurate single method on the full cohort at every horizon (MAE 3.02*/*1.99*/*1.52 at 3/6/9 months). Paired tests confirm carry-forward significantly outperforms the LLM on the full cohort (LLM*−*History ΔMAE *−*0.054*/ −* 0.071*/ −* 0.075; Wilcoxon *p* = 0.046*/*0.001*/*0.0006). We state this plainly: *any* model claiming to forecast PASC severity must be judged against carry-forward, and on stable patients it is the appropriate choice.

### 3.2 Among learned models, the LLM regressor is best

Among the *learned* predictors (Lasso, Ridge, XGBoost, MLP, ResNet, Attention) the LLM regressor had the lowest MAE in every cell of Table 2. The gap over the strongest tree model is real: at 3 months LLM MAE 3.11 vs. XGBoost 3.57, with paired LLM-vs-XGBoost *p ≤* 0.01 on the full, responder, and protected cohorts. On squared error the LLM additionally beats carry-forward on the full cohort at the longer horizons (e.g. 6-month MSE 14.6 vs. History 16.7), indicating it reduces large errors even where it loses on median error—a desirable property when the cost of a large mis-estimate is high.

### 3.3 Where learning helps: the dynamic Responder phenotype

The phenotype stratification (Table 2, right; Fig. 2) localizes the benefit. In the **Responder** group—patients whose trajectories actually move—the LLM is the most accurate method *overall*, beating even carry-forward at 3 months (MAE 4.72 vs. 4.82) and 6 months (4.06 vs. 4.36). We are careful not to over-claim: the LLM-vs-carry-forward difference here is *not* statistically significant (*p* = 0.70 at 3m, 0.29 at 6m; *n* = 10 resamples, Responder *N* = 349*/*124*/*88), and at 9 months the two tie. This is a hypothesis-generating signal consistent with intuition: when the last observation is a poor proxy for the future (dynamic trajectories), a model that integrates physiology and history adds value; when trajectories are flat (Protected), nothing beats carry-forward.

**Figure 2:**
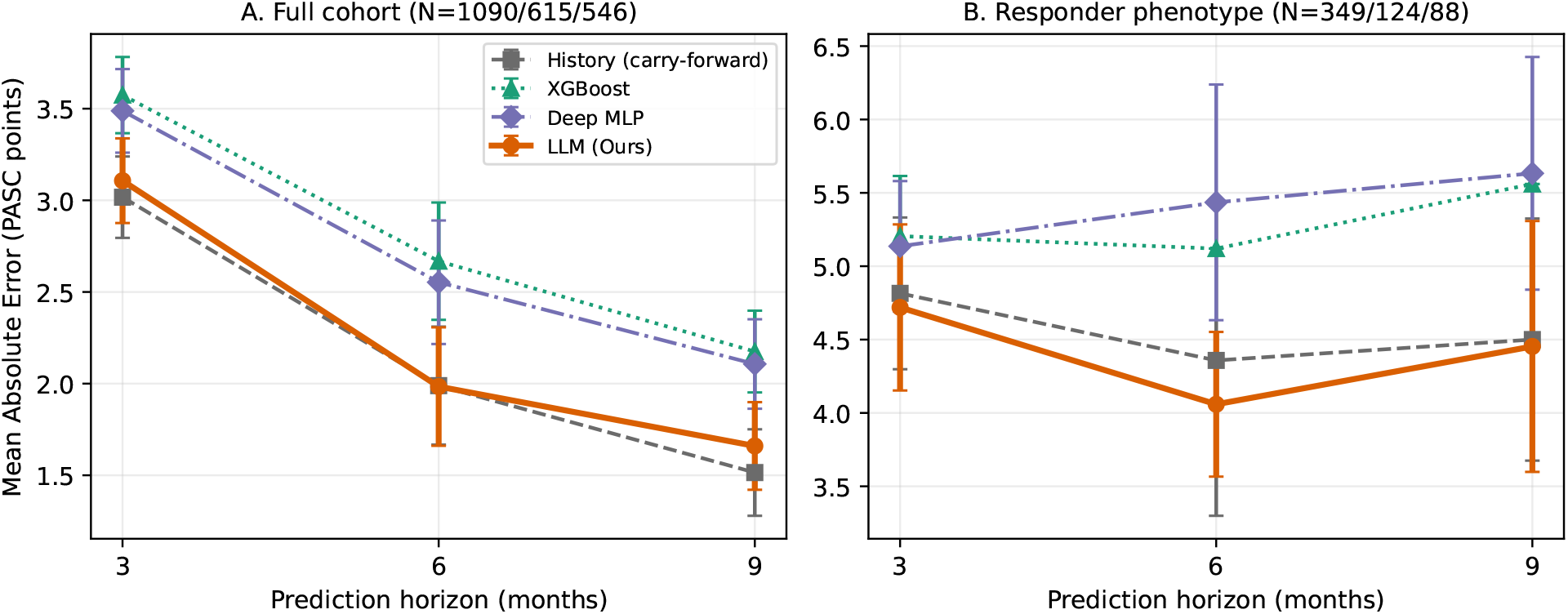
Mean absolute error versus prediction horizon (error bars: SD over resamples). **(A)** Full cohort: carry-forward (History) is the reference and is hardest to beat; the LLM regressor tracks it closely and leads the other learned models. **(B)** Responder phenotype: the LLM is the most accurate method at 3 and 6 months (advantage over carry-forward not statistically significant). XGBoost SD omitted where a single configuration was evaluated.

### 3.4 Interpretability: the model suppresses confounders and keeps pathology

The central claim of the paper is interpretive, not metric. We extracted per-token saliency from the disentanglement attention (the gate routing each token toward the causal head). The pattern (Table 3, Fig. 3), obtained from the disentanglement variant at the 3-month horizon, is clinically sensible: direct pathology tokens (*breathlessness, malaise, unrefreshing sleep*) reach maximal saliency (1.00); neuro-PASC tokens (*brain fog, insomnia*) are intermediate (0.54–0.74); and the principal confounders—*menopause* and *diabetes*—are suppressed (< 0.27), as is linguistic filler (< 0.17). In other words the model learns to predict future severity from disease semiotics while down-weighting the very comorbidity/transition tokens that would otherwise drive over-diagnosis in this population.

**Figure 3:**
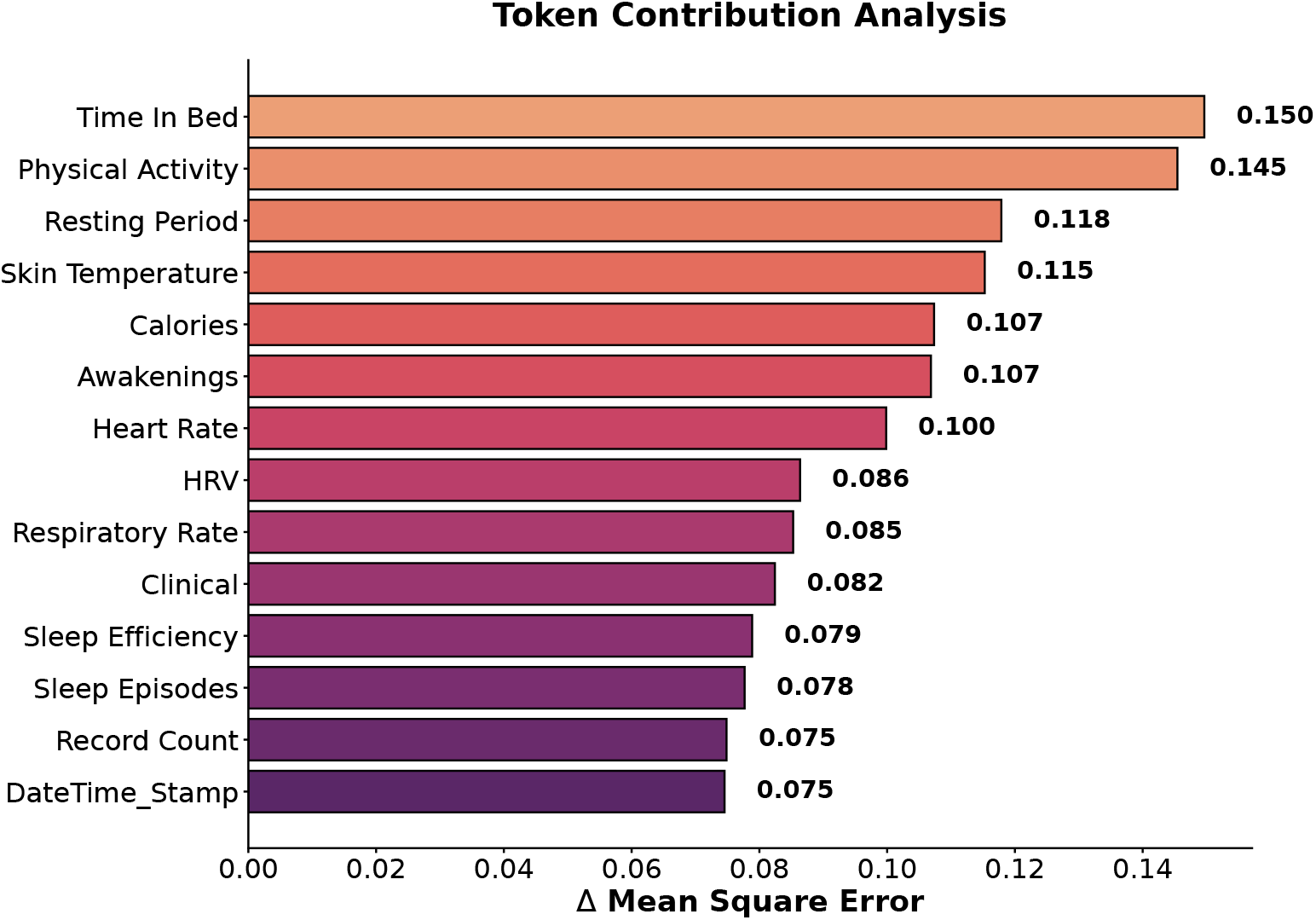
Token-level saliency from the disentanglement layer. Pathology tokens dominate the causal head while confounders (menopause, diabetes) and filler are suppressed.

## 4 Discussion

We set out to forecast Long-COVID severity in women while resisting the population-specific hazard that menopause and comorbidities are confounded with PASC symptomatology. Three findings stand out, and we present them in the order of their honesty rather than their flattery.

First, **carry-forward is the reference and it is strong**. PASC severity is autocorrelated, so predicting that a patient’s next score equals her last is hard to beat on a cohort dominated by stable trajectories. We report this as the primary benchmark, contrary to a common temptation to compare only against weak learned baselines. Any future PASC predictor should be evaluated this way.

Second, **the value of learning is conditional and modest on accuracy**. Among learned models the LLM regressor is uniformly best and significantly beats gradient-boosted trees; on squared error it beats carry-forward at longer horizons; and in the dynamic Responder phenotype it is the most accurate method overall—though that final, clinically interesting advantage is not statistically significant at our sample size. We therefore frame the Responder result as hypothesis-generating and explicitly power-limited.

Third, and most important, **the model’s contribution is interpretive**. The disentanglement variant’s attention assigns maximal weight to pathology and suppresses menopause and diabetes. No error metric rewards this, yet it is precisely what a deployable clinical tool in a confounded, women’s-health setting needs: a “clinically honest” attribution that resists misattributing baseline physiology to viral disease. A slightly less accurate but causally transparent model can be preferable to a marginally more accurate but opaque one when the failure mode is systematic over-flagging of a large subpopulation.

### Limitations

(1) Single program (RECOVER); we report internal resampling and phenotype stratification but no external-cohort validation. (2) Cohort attrition grows with horizon and the Responder subgroup is small (*N* down to 88 at 9 months), limiting power—hence the non-significant subgroup result. (3) For longer horizons the wearable window overlaps the outcome survey, so those inputs are monitoring rather than strictly pre-baseline features; a baseline-only sensitivity analysis is planned. (4) Wearable coverage is partial and handled by sentinel imputation. (5) The backbone is a small (0.5B) model; larger encoders may shift the accuracy picture. (6) Phenotype thresholds are inherited from the RECOVER extraction and not re-derived here. (7) The PASC index is self-report–based. (8) Two model configurations share one backbone: the multi-horizon accuracy results (Table 2) use the residual-fusion LLM regressor, whereas the saliency/attribution results (Table 3) use the causal-disentanglement variant, which we evaluated at the 3-month horizon. We therefore do not claim the disentanglement variant matches the regressor’s multi-horizon error, and the multi-horizon stability of the attribution pattern is reported as preliminary. Running the disentanglement variant at all horizons—so a single model carries both the accuracy and the interpretability story—is the primary planned extension.

### Future work

External validation (e.g. a second RECOVER site or an independent wearable cohort), a strictly baseline-only predictive variant, calibration and decision-curve analysis for the Responder-flagging use case, and a formal ablation isolating each disentanglement component’s contribution to confounder suppression.

### Conclusion

For Long-COVID severity forecasting in women, last-value carry-forward is a strong and honest reference; the contribution of a learned, causally disentangled language model is better accuracy where trajectories are dynamic and, above all, an interpretable attribution that down-weights confounders such as menopause. This positions the approach as a transparency-enhancing complement to, rather than a wholesale replacement for, simple clinical heuristics.

## Data availability

Individual-level RECOVER data are available to qualified researchers through the NIH RECOVER data program (https://recovercovid.org/data) under its data-use terms; the authors cannot redistribute participant-level data. Analysis code and the trajectory-phenotype definitions will be made available in a public repository upon publication.

## Author contributions

J.W. and J.C.W. conceived the study and clinical framing. J.W., J.S., and T.Z. designed the model and experiments. J.S. implemented the multi-horizon pipeline and baselines. Y.L. and A.S. provided clinical interpretation. X.N. and Q.X. advised on methodology and statistics. Z.G. advised on the women’s-health and cardiopulmonary framing. J.W. drafted the manuscript; all authors revised and approved it.

## Acknowledgments

This research was supported by the NIH Intramural Research Program, National Library of Medicine, and by the NIH RECOVER Initiative. The views are those of the authors and do not necessarily represent those of the NIH or the U.S. Government. The authors thank Dean Foster (Amazon) for valuable insights and discussions that helped shape this work, and Renee Sears (Velsera) for platform support.

## Footnotes

1 https://recovercovid.org/data

## References

[1] National Academies of Sciences, Engineering, and Medicine. A long COVID definition: a chronic, systemic disease state with profound consequences. The National Academies Press, Washington, DC, 2024.

[2] Tanayott Thaweethai, Sarah E Jolley, Elizabeth W Karlson, Emily B Levitan, Bruce Levy, Grace A McComsey, Lisa McCorkell, Girish N Nadkarni, Sairam Parthasarathy, Upinder Singh, et al. Development of a definition of postacute sequelae of sars-cov-2 infection. Jama, 329(22):1934–1946, 2023.

[3] Hannah E Davis, Lisa McCorkell, Julia Moore Vogel, and Eric J Topol. Long covid: major findings, mechanisms and recommendations. Nature Reviews Microbiology, 21(3):133–146, 2023.

[4] Dimpy P Shah, Tanayott Thaweethai, Elizabeth W Karlson, Hector Bonilla, Benjamin D Horne, Janet M Mullington, Juan P Wisnivesky, Mady Hornig, Daniel J Shinnick, Jonathan D Klein, et al. Sex differences in long covid. JAMA network open, 8(1):e2455430, 2025.

[5] Lars G Fritsche, Weijia Jin, Andrew J Admon, and Bhramar Mukherjee. Characterizing and predicting post-acute sequelae of sars cov-2 infection (pasc) in a large academic medical center in the us. Journal of Clinical Medicine, 12(4):1328, 2023.

[6] Zahin Amin-Chowdhury and Shamez N Ladhani. Causation or confounding: why controls are critical for characterizing long covid. Nature Medicine, 27(7):1129–1130, 2021.

[7] Miguel A Hernán and James M Robins. Causal inference. CRC Boca Raton, FL, 2010.

[8] Erik Igelström, Peter Craig, Jim Lewsey, John Lynch, Anna Pearce, and Srinivasa Vittal Katikireddi. Causal inference and effect estimation using observational data. J Epidemiol Community Health, 76(11):960–966, 2022.

[9] Laura D Straus, Xinming An, Yinyao Ji, Samuel A McLean, Thomas C Neylan, AURORA Study Group, Ayse S Cakmak, Anne Richards, Gari D Clifford, Mochuan Liu, et al. Utility of wrist-wearable data for assessing pain, sleep, and anxiety outcomes after traumatic stress exposure. JAMA psychiatry, 80(3):220–229, 2023.

[10] Aitolkyn Baigutanova, Sungkyu Park, Marios Constantinides, Sang Won Lee, Daniele Quercia, and Meeyoung Cha. A continuous real-world dataset comprising wearable-based heart rate variability alongside sleep diaries. Scientific data, 12(1):1474, 2025.

[11] Michael Moor, Oishi Banerjee, Zahra Shakeri Hossein Abad, Harlan M Krumholz, Jure Leskovec, Eric J Topol, and Pranav Rajpurkar. Foundation models for generalist medical artificial intelligence. Nature, 616(7956):259–265, 2023.

[12] Zilong Bai, Zihan Xu, Cong Sun, Chengxi Zang, H Timothy Bunnell, Catherine Sinfield, Jacqueline Rutter, Aaron Thomas Martinez, L Charles Bailey, Mark Weiner, et al. Extracting post-acute sequelae of sars-cov-2 infection symptoms from clinical notes via hybrid natural language processing. npj health systems, 2(1):31, 2025.

[13] Binyuan Hui, Jian Yang, Zeyu Cui, Jiaxi Yang, Dayiheng Liu, Lei Zhang, Tianyu Liu, Jiajun Zhang, Bowen Yu, Keming Lu, et al. Qwen2. 5-coder technical report. arXiv preprint 2409.12186, 2024.

[14] Advait Parulekar, Liam Collins, Karthikeyan Shanmugam, Aryan Mokhtari, and Sanjay Shakkottai. Infonce loss provably learns cluster-preserving representations. In The thirty sixth annual conference on learning theory, pages 1914–1961. PMLR, 2023.

[15] Robert Tibshirani. Regression shrinkage and selection via the lasso. Journal of the Royal Statistical Society Series B: Statistical Methodology, 58(1):267–288, 1996.

[16] Gary C McDonald. Ridge regression. Wiley Interdisciplinary Reviews: Computational Statistics, 1(1):93–100, 2009.

[17] Tianqi Chen and Carlos Guestrin. Xgboost: A scalable tree boosting system. In International Conference on Knowledge Discovery and Data mining, pages 785–794, 2016.

[18] Kaiming He, Xiangyu Zhang, Shaoqing Ren, and Jian Sun. Deep residual learning for image recognition. In Proceedings of the IEEE conference on computer vision and pattern recognition, pages 770–778, 2016.

[19] Ashish Vaswani, Noam Shazeer, Niki Parmar, Jakob Uszkoreit, Llion Jones, Aidan N Gomez, L ukasz Kaiser, and Illia Polosukhin. Attention is all you need. Advances in neural information processing systems, 30, 2017.

